# Effectiveness of AI-Supported Holistic Health Management Through PREVERY’s Digital Health Promotion and Prevention Program: A Prospective Before-and-After Observational Study

**DOI:** 10.64898/2026.11.07.26367492

**Authors:** Miriam Ringleb, Timo Trampert, Tamira Trampert, Wilhelm Bloch

**Affiliations:** NeuroPyschoImmunology research unit, Department for Molecular and Cellular Sports Medicine, Institute of Cardiovascular Research and Sports Medicine, German Sport University Cologne, Cologne, Germany; Center for Interdisciplinary Prevention of Diseases related to Professional Activities, Friedrich-Schiller-University Jena, Jena, Germany; Department of Sports Medicine and Health Promotion, Friedrich-Schiller-University Jena, Jena, Germany; PREVERY UG, Cologne, Germany; Department for Molecular and Cellular Sports Medicine, Institute of Cardiovascular Research and Sports Medicine, German Sport University Cologne, Cologne, Germany

## Abstract

**Background:** There is an increasing strain on our healthcare system due to rising numbers of diseases related to chronic stress. One approach to combat these stress-related complaints is health coaching. However, personal health coaching is expensive and time costly, while digital offers can complement the already existing health coaching strategies. Therefore, this study investigates the effects of an AI-supported, digital health and prevention program.

**Methods:** 253 participants were included in the study, with 113 being analyzed eventually. General stress experience and self-efficacy were assessed at baseline (T1), after six weeks (T2), and three months after the interventional phase (T3) via a Numerical Analog Scale (NAS) with values ranging from 0 to 10, the Perceived Stress Scale (PSS), and the PsyCap Scale (CPC).

**Results:** A significant time effect was observed for CPC and PSS. CPC increased significantly from T1 to T2 and T3 and remained stable between T2 and T3, while PSS decreased from T1 to T2 and increased again by T3. All pairwise comparisons were significant. There were also significant time effects for self-reported current health status, health literacy, health awareness, and self-efficacy, with values at T3 being higher than at T1 (for health awareness, this was already evident at T2; for self-efficacy, this was additionally evident between T2 and T3), while no time effect was found for motivation.

**Conclusion:** PREVERY seems to be an effective prevention program to increase psychological capital together with self-reported current health status, health awareness, health literacy, and self-efficacy, while at the same time reducing perceived stress.

## 1 Introduction

Stress-related complaints are widespread and associated with significant health and economic consequences. According to the WHO (2022), the global economy loses 12 billion workdays each year, resulting in costs of approximately 1 trillion U.S. dollars, due to stress, depression, and anxiety. Especially, coupled with low levels of resilience and self-efficacy, chronic stress can have serious mental and physical consequences, such as depression (Tafet & Bernardini, 2003), sleep disorders (Lo Martire et al., 2020), an increased risk of cardiac events (Dimsdale, 2008), and a rise in chronic low-grade inflammation (Rohleder, 2019). Since persistent psychological strain poses a major threat to public health, effective and scalable solutions are needed to meet the demand for stress management measures. As stress-related chronic diseases are largely preventable, the importance of lifestyle interventions in rehabilitation and prevention is undisputed (Hendrickx et al., 2005; Warburton, 2006).

Taking a more proactive approach to raise awareness for the own health and changing one’s health behaviors is, therefore, essential to reduce the burden on the healthcare system. One promising approach supporting the implementation of health behavioral changes is the use of health coaching (Dejonghe et al., 2017). It has been shown that health coaching can help to attain a specific diet (e.g., in a diabetes context), increase physical activity, or reduce body mass index (BMI) in overweight individuals (Olsen & Nesbitt, 2010). Additionally, it was found that health coaching can lower depressive symptoms (Sacco et al., 2009), improve self-efficacy (Linden et al., 2010), and reduce stress significantly (Wolever et al., 2010).

However, most interventions use a face-to-face set-up to conduct their health coaching, which is expensive and there is limited access for the overall population to take advantage of this offer. For this reason, innovative strategies are highly relevant in providing an effective and scalable alternative to complement existing solutions (Loughnane et al., 2025). Digital, behavior-oriented programs targeting the change in health behavior of the overall population are one possible approach. They may reduce the experience of stress, particularly when they combine knowledge, goal-setting, self-reflection, motivational strategies, and practical everyday exercises.

A study conducted by Ong et al. (2024) found that large language models (LLM) performed comparably to health coaches across five quality dimensions and aligned with experts’ assessments in terms of accuracy and probability of harm. In 60% of cases, the AI-generated responses were even preferred over those provided by real coaches. Additionally, chatbot-delivered coaching apps were able to reduce stress (Ulrich et al., 2024) and increase goal attainment (Terblanche et al., 2022). Although these positive effects of mobile health (mHealth) interventions are known, evidence on LLM-based coaching embedded in a structured prevention program is lacking, particularly in German-speaking populations.

Based on this, the PREVERY health counseling app was developed to address this gap in innovation. The digital prevention platform supports users through an artificial intelligence (AI)-based health coach (EVE) in promoting health and, in particular, stress and resource management. In addition to perceived stress, psychological capital was assessed as a broader construct comprising self-efficacy, hope, resilience, and optimism (Luthans et al., 2007). Within this concept, self-efficacy is an independent construct (D. M. Williams & Rhodes, 2016). Being under high stress or chronic strain, increases the risk that the psychological resources will be depleted. Conversely, high self-efficacy and resilience act as a buffer against stress by helping to perceive stressors as manageable challenges rather than threats. Therefore, stress and psychological capital, together with self-efficacy, can serve as primary direct targets in developmental interventions (Corbu et al., 2021).

In general, PREVERY’s approach is based on especially two established scientific models and principles in the field of health-promoting behavioral psychology. The first one is the Health Action Process Approach (HAPA)-model (Schwarzer, 2004). It is a social–cognitive model specifying motivational and volitional determinants of health behavior. The HAPA-model assumes that people first form an intention and then develop strategies to translate that intention into concrete action (Schwarzer, 2004). The model distinguishes between a motivational phase (decision-making and goal-setting) and a volitional phase (planning, implementation, and maintenance). Three factors are crucial here: self-efficacy, resources, and coping with barriers (Schwarzer, 2004). PREVERY makes targeted use of this principle: During the goal-setting process, resources and potential barriers are reflected upon and identified to promote realistic and sustainable action plans, thereby systematically supporting motivation, planning, and implementation.

The second model is the biopsychosocial model developed by Engel (1980). According to this model, health and illness arise from the interplay of biological, psychological, and social factors. None of these dimensions exists in isolation as they influence one another and collectively determine how to feel, think, and act (Lehman et al., 2017). For example, stress or social support affect physical health just as much as exercise or nutrition affect mental health. PREVERY is guided precisely by this understanding, addressing six areas of health (body and exercise, nutrition and enjoyment, stress and recovery, mind and emotions, meaning and quality of life, and environment and social life) based on the three dimensions of the model.

Generally, PREVERY can best be compared to the analogous process of professionally conducted systemic health coaching, which has been examined in comparable studies (Dejonghe et al., 2017). It systematically addresses the following mechanisms: onboarding, comprehensive self-assessment across six health domains (body and movement, diet, stress and recovery, mind and emotion, quality of life, environment), personalized results reports, SMART (specific, measurable, feasible, realistic, scheduled) goals, behavior-based tracking with gamification, and guided reflection dialogs with the AI coach EVE.

The aim of this study is to determine whether such an analog process, by embedding generative AI in the form of a LLM into a guided coaching process, offers a meaningful and scalable alternative to an in-person health coaching. The primary endpoint is the general stress experience. Secondary endpoints include the psychological capital, self-reported health status, self-efficacy, health literacy and awareness, and motivation. The intervention lasts six weeks, with assessments at baseline (T1), after six weeks (T2), and at three months follow-up (T3). It is hypothesized, that the use of the PREVERY app will lead to reductions in stress levels and improvements in the psychological capital, self-reported health status, self-efficacy, health literacy and awareness, and motivation from T1 to T2 and T3. The goal is to demonstrate the program’s effectiveness in accordance with the requirements of the Central Review Board for Prevention (ZPP) so that individuals with statutory health insurance can be reimbursed for the costs of the prevention program following certification, as well as to evaluate the PREVERY app as a scalable, AI-powered tool for health promotion in the area of stress and resource management.

## 2 Methods and materials

This efficacy study is a collaborative project between the digital health company PREVERY UG and BERGISCHE KRANKENKASSE, with the participation of the Institute for Cardiovascular Research and Sports Medicine German Sport University Cologne. The study has been registered in the German Clinical Trials Register (DRKS00037999). Ethics approval was obtained by the Ethics Committee of the German Sport University Cologne on 25 September 2025 (reference number: 197/2025).

### 2.1 Experimental design

This is a prospective before-and-after observational study (single-group pre-post design). Participants had to use the digital health program PREVERY for six weeks. In addition, the participants were required to complete a questionnaire at three different time points (before the start, after six weeks of active app use, and after three months). Self-assessments of current health, health awareness, health literacy, self-efficacy, and motivation were measured using a Numerical Analog Scale (NAS) with values ranging from 0 to 10. In addition, the Perceived Stress Scale, which includes subscales for helplessness and self-efficacy (to measure perceived stress levels), was administered to assess “changes in overall stress experience”, along with the PsyCap Scale.

#### 2.1.1 App content and usage

After onboarding, users completed a holistic self-assessment. The Health Compass is created based on holistic questions about well-being. The user then selects a focus area. Within this focus area, the user evaluates statements regarding official behavioral recommendations (e.g., 150 minutes of moderate-intensity endurance exercise) based on how they are actually implemented in daily life. The user then selects one of the lower-rated and more generic behavioral recommendations to develop a SMART goal from it through a guided process. Based on this, users receive personalized health impulses, complete app-based exercises, track their behavior, and reflect on their progress with the AI-supported health coach EVE. The app combines elements of health education, behavioral psychology, goal setting, self-reflection, gamification, and guided digital coaching.

Generally, a simple interface to OpenAI’s ChatGPT was used (Chat-GPT_Modell 4o). An opt-out provision prevented the models from being trained using the participants’ data only. Otherwise, it was ensured that data processing during the pilot complied with the GDPR. Highly restrictive prompts and a predefined evidence-based knowledge base were used to minimize the risk of AI-generated hallucinations and ensure that responses remained aligned with the structured prevention process. The AI relied primarily on user inputs and the well-researched database as its “source of truth”. There was no human oversight of the AI’s output. There was no possibility of a free chat communication between the participants the AI. Participants had the option to consult the study team at any time.

For the purpose of the study, a predefined minimum usage criterion was applied. Participants were required to collect at least 20 growth points in each intervention week. Growth points could be obtained by completing different app-based tasks, including reflection exercises, knowledge units, goal-related activities, tracking tasks, and interactions within the guided coaching process. Based on the structure and expected duration of these tasks, 20 growth points corresponded approximately to a minimum app usage of 30 minutes per week. A more detailed description on the amount of growth points given per completed task can be found in the Supplementary Material. It must be noted that the implementation of the set goals took place in addition to the 30 minutes and on an individual basis as part of daily life. This minimum usage criterion was implemented to operationalize a measurable level of intervention exposure in line with the requirements for evaluating digital prevention and health promotion programs. Participants who did not reach the minimum of 20 growth points in each of the six intervention weeks were not included in the final per-protocol analysis.

#### 2.1.2 Survey and Measurement Tools

A mixed-methods approach using questionnaires was employed. The validated measures used were the Psychological Capital Scale (PsyCap Scale (CPC-12R)) for the primary endpoint psychological capital, and the Perceived Stress Scale (PSS-10, German version) for the “change in general stress experience”. The PsyCap Scale (CPC-12R) is a 12-item, revised version of the Compound Psychological Capital Scale, which measures an individual’s psychological capital (PsyCap). It assesses the four components of PsyCap - self-efficacy, hope, resilience, and optimism - with three items each. The CPC-12R has been validated in English- and German-speaking samples and demonstrates reliability, validity, and measurement invariance across cultures (Dudasova et al., 2021).

The PSS developed by Cohen et al. (1983) is a widely used and well-established self-report scale for measuring perceived stress. It comprises two subscales: perceived helplessness (items 1, 2, 3, 6, 9, 10), which captures the feeling of a lack of control over circumstances, emotions, or reactions, and lack of self-efficacy (items 4, 5, 7, 8), which measures the subjective assessment of one’s own problem-solving ability. The total score is calculated as the sum of the items on the helplessness scale and the sum of the inverted items on the self-efficacy scale, allowing both primary outcomes to be assessed using the PSS. As part of a representative study, the PSS-10 was psychometrically evaluated, and normative values were established for the German population. The results demonstrated good internal consistency and construct validity. Perceived stress was consistently associated with depression, anxiety, exhaustion, procrastination, and reduced life satisfaction. The PSS-10 thus proved to be a reliable, valid, and economical instrument for measuring stress (Klein et al., 2016).

#### 2.1.3 Eligibility criteria

Inclusion criteria were a minimum age of 18 years, the willingness to use the PREVERY app (language: German) for six weeks, to have access to an internet-enabled device, and the consent to participate in the study. Exclusion criteria were acute psychological diseases, severe eating disorders, and untreated chronic conditions without medical supervision.

### 2.2 Sample size calculation

The sample size calculation is based on an a priori power analysis using a repeated measures ANOVA (pre-post comparison within the same group). Assuming a significance level of α = 0.05 and a target statistical power of 1–β = 0.80, the calculation yielded a required minimum sample size of n = 34 participants. To account for potential dropouts (a dropout rate of 20%), the target sample size was adjusted accordingly. This results in a planned target sample size of n = 43 participants to ensure the study’s statistical power. The selected effect size is based on existing studies on digital prevention measures for stress reduction and the promotion of self-efficacy, in which moderate to large effects were reported (Gong et al., 2023; Heber et al., 2017). However, if a smaller effect (e.g., *d* = 0.20) is assumed - as might occur in real-world settings or with low intervention intensities - up to n = 249 participants would be required. Against this backdrop, aiming for a sample size of approximately 150 participants is a sensible, conservative approach to ensure that even smaller effects can be detected, to make the results more robust and generalizable, and to allow for subgroup analyses (e.g., by sex, age, or usage intensity) if necessary.

### 2.3 Statistics

All analysis were conducted in R (version 2026.09.0). Participants with data at all three time points (T1, T2, T3) were included in the analysis (*n* = 113). Each outcome (PSS and PSC scores, current health, health awareness, health literacy, self-efficacy, and motivation) was analyzed separately. Descriptive statistics are reported as means, standard deviations, and medians for each time point. Distribution was tested via skewness, kurtosis, histograms, Q-Q-plots, and Shapiro-Wilk test. Although the Shapiro-Wilk test was significant for PSS and CPC, visual inspection of the Q-Q-plots still allows for the use of parametric tests. Repeated measures ANOVAs were computed for all outcomes, with Greenhouse-Geisser correction applied when Mauchly’s test indicated a violation of sphericity (generalized eta squared, η², as effect size) and Bonferroni-adjusted paired *t*-tests as post hoc tests. Effect sizes (comparable to Cohen’s *d*) for the pairwise comparisons are reported as the mean difference divided by the average of the standard deviation at the two time points. For CPC, two values were 3z winsorization, while no correction was needed for PSS. As data for current health, health awareness, health literacy, self-efficacy, and motivation were ordinally distributed, leading to skewed and not normally distributed distributions, Friedman tests were conducted as sensitivity analyses with Kendall’s *W* as effect size. Significant Friedman tests were followed by pairwise Wilcoxon signed-rank tests, with *p* values adjusted using the Bonferroni method; effect sizes for pairwise comparisons are reported as *r*. No winsorization was performed for these outcomes as extreme values were plausible responses within the scale range and were therefore retained. The level of significance was set at α = .05 (two-sided).

## 3 Results

### 3.1 Participant characteristics

Two hundred-fifty-three participants were included in the study. Most dropouts (n = 139) had to be excluded as they did not meet the minimum usage time of at least 20 growth points in each intervention week (Figure 1). One additional dropout was due to not fulfilling the questionnaires at T3. Therefore, 113 participants (77 females, 35 males, 1 divers) with an average age of 47±13 years were included in the analysis. The participants who completed the full six weeks obtained on average 56±12 growth points per week.

**Figure 1.**
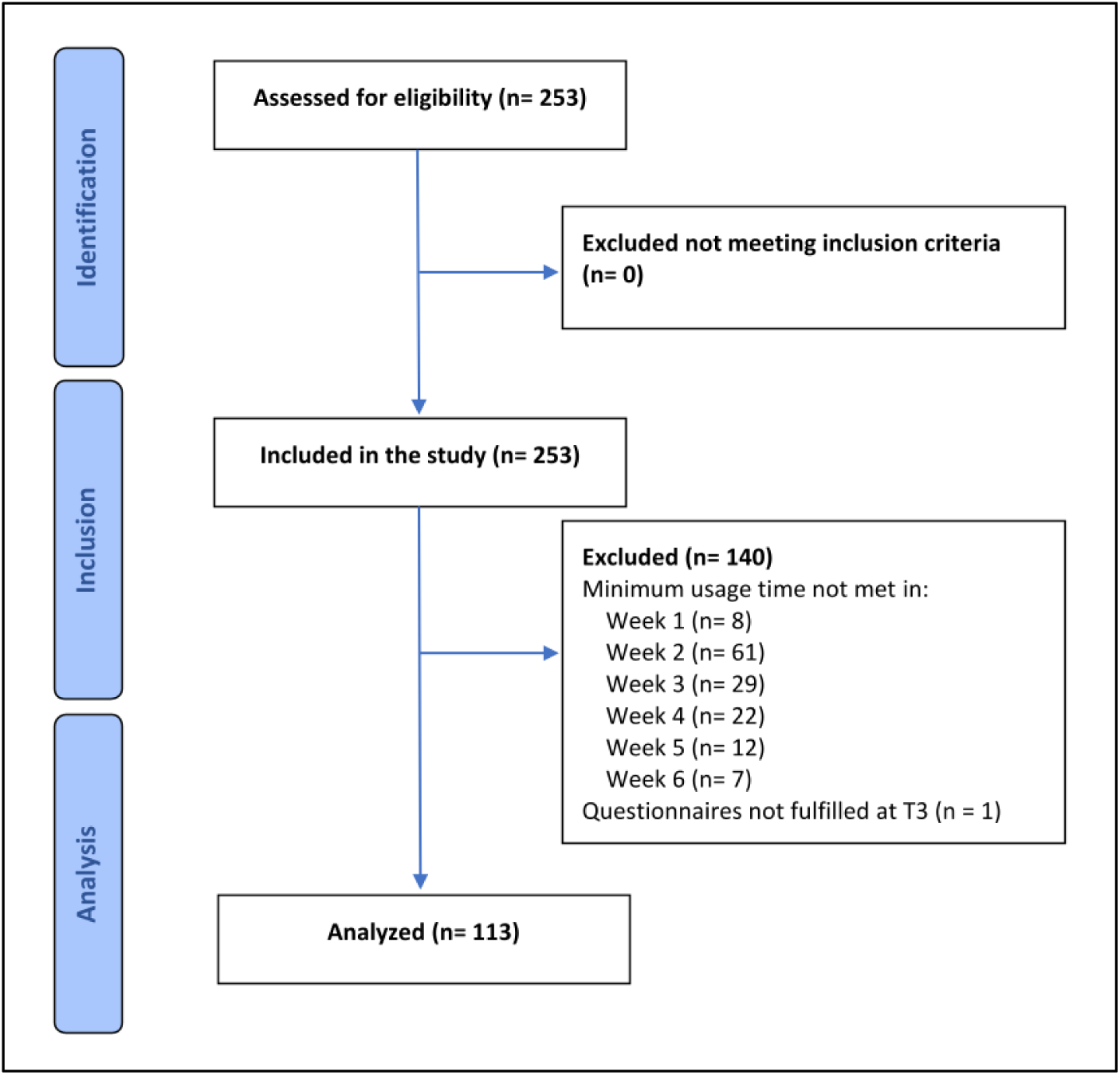
Participant flowchart

### 3.2 Psychological capital and perceived stress

Results for PSS and CPC are displayed in Figure 2 and effect sizes in Table 1. For both CPC and PSS, Mauchly’s test indicated a violation of the sphericity assumptions, which is why Greenhouse-Geisser-corrected degrees of freedom are reported.

**Figure 2.**
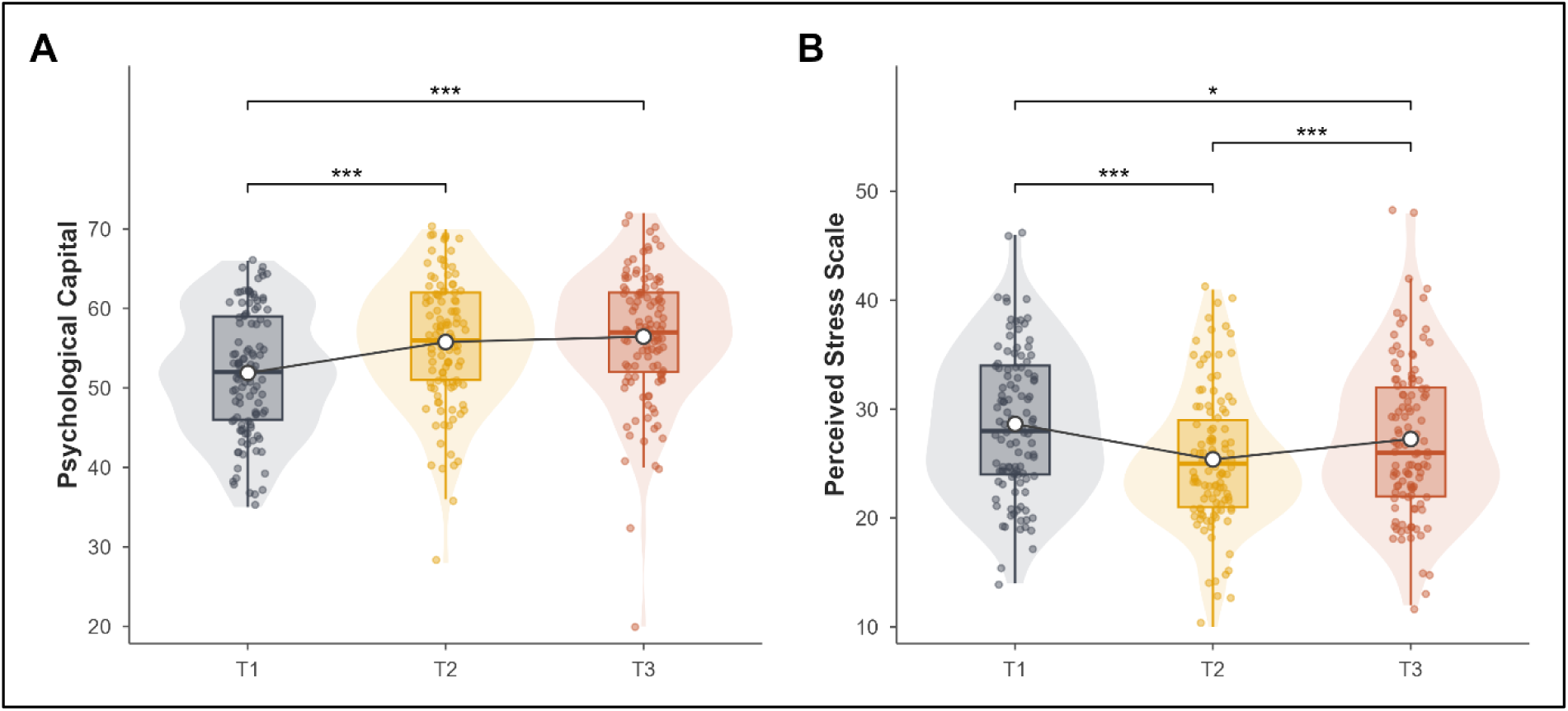
Psychological capital (A) and perceived stress (B) at three measurement time points (T1–T3; n = 113). The violin plots show the distribution of values, the box plots indicate the median and interquartile range (whiskers: 1.5 × IQR), and the points represent individual participants. White circles connected by a line indicate the mean at each time point. Parentheses indicate significant pairwise differences between time points (paired t-tests, Bonferroni-corrected following repeated-measures ANOVA): * p < 0.05, *** p < 0.001.

**Table 1.** Effect sizes for the outcomes psychological capital and perceived stress.

| Outcome | Comparison | Effect size (*d*) |
|---|---|---|
| **Psychological capital** | **T1 vs. T2** | **0.70\*\*\*** |
|  | **T1 vs. T3** | **0.81\*\*\*** |
|  | T2 vs. T3 | 0.19 |
| **Perceived Stress** | **T1 vs. T2** | **-0.63\*\*\*** |
|  | **T1 vs. T3** | **-0.25\*\*\*** |
|  | **T2 vs. T3** | **0.45\*\*\*** |

*Note* T1 represents levels pre-intervention, T2 directly post-intervention, and T3 three months follow-up. Significant pairwise differences between time points (paired t-tests, Bonferroni-corrected following repeated-measures ANOVA) are indicated as follows: \*\*\* p < 0.001.

There was a significant time effect for CPC (*F*(1.69, 189.82) = 52.88, *p* < .001, *η²* = .065). Bonferroni-corrected post hoc comparisons showed that the values differed significantly between T1 and T2, *t*(112) = 5.28, *padj* < 0.001, *d* = 0.70, as well as between T1 and T3, *t*(112) = 6.64, *padj* < 0.001, *d* = 0.81. There was no significant difference between T2 and T3, *t*(112) = 1.10, *padj* = 0.819, *d* = 0.19.

Additionally, there was a significant time effect for PSS (*F*(1.80, 201.45) = 23.88, *p* < .001, *η²* = .040). Bonferroni-corrected post hoc comparisons revealed significant differences between all time points: T1 vs. T2, *t*(112) = -26.20, *padj* < 0.001, *d* = -0.63; T1 vs. T3, *t*(112) = -17.00, *padj* < 0.001, *d* = -0.25, and T2 vs. T3, *t*(112) = 14.10, *padj* < 0.001, *d* = 0.45.

### 3.3 Current health, health awareness, health literacy, self-efficacy, and motivation

Results for self-reported current health, health awareness, health literacy, self-efficacy, and motivation are displayed in Figure 3 and effect sizes in Table 2.

**Figure 3.**
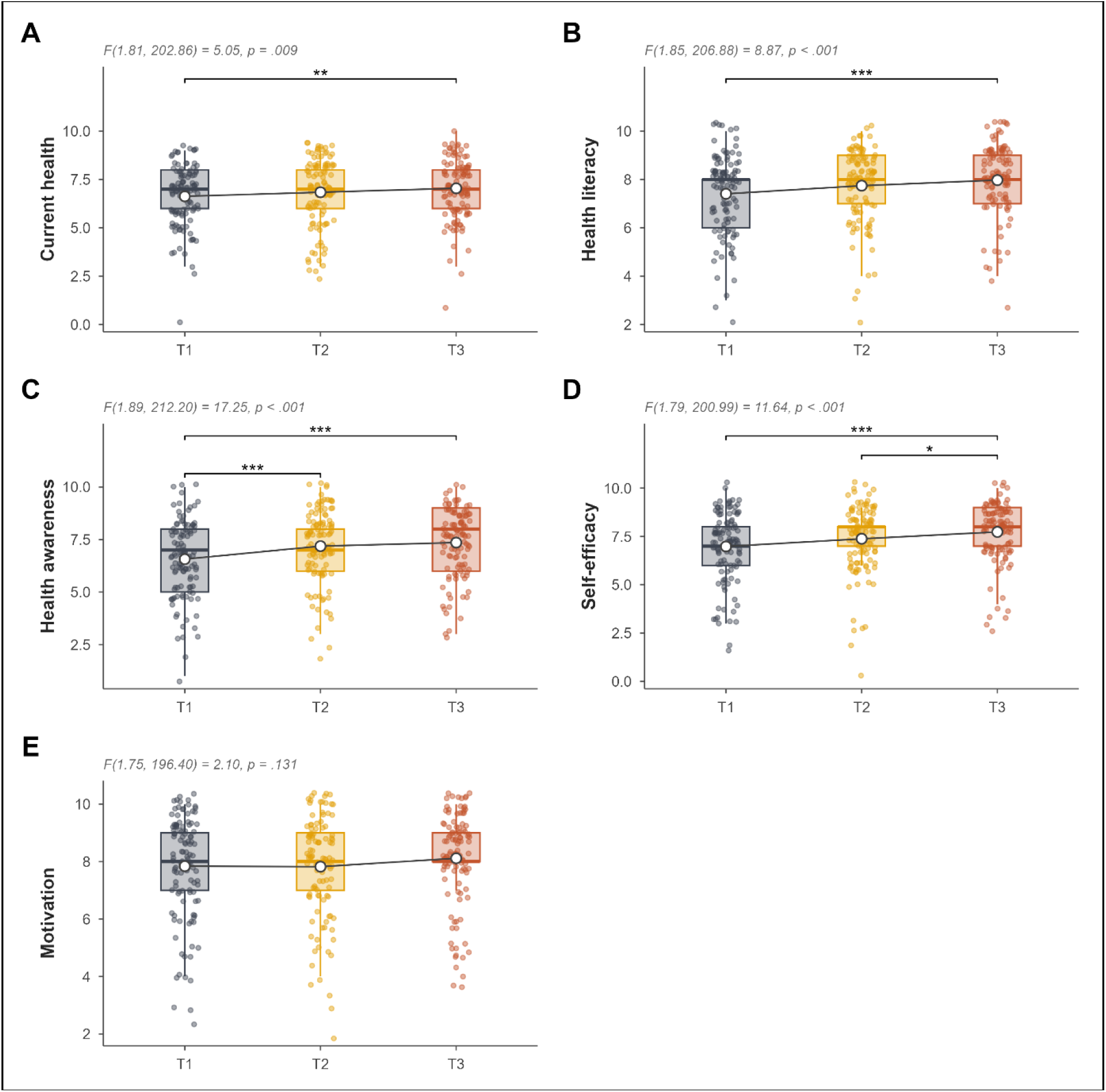
Current health status (A), health literacy (B), health awareness (C), self-efficacy (D), and motivation (E) at the three time points (T1–T3; *n* = 113). All items were rated on a scale from 0 to 10. The violin plots show the distribution of values, the box plots indicate the median and interquartile range (whiskers: 1.5 × IQR), and the points represent individual participants (jittering for better visibility). White circles connected by a line indicate the mean at each time point. Parentheses indicate significant pairwise differences between time points (paired t-tests, Bonferroni-corrected following repeated-measures ANOVA): * p < 0.05, ** p < 0.01, *** p < 0.001.

**Table 2.**
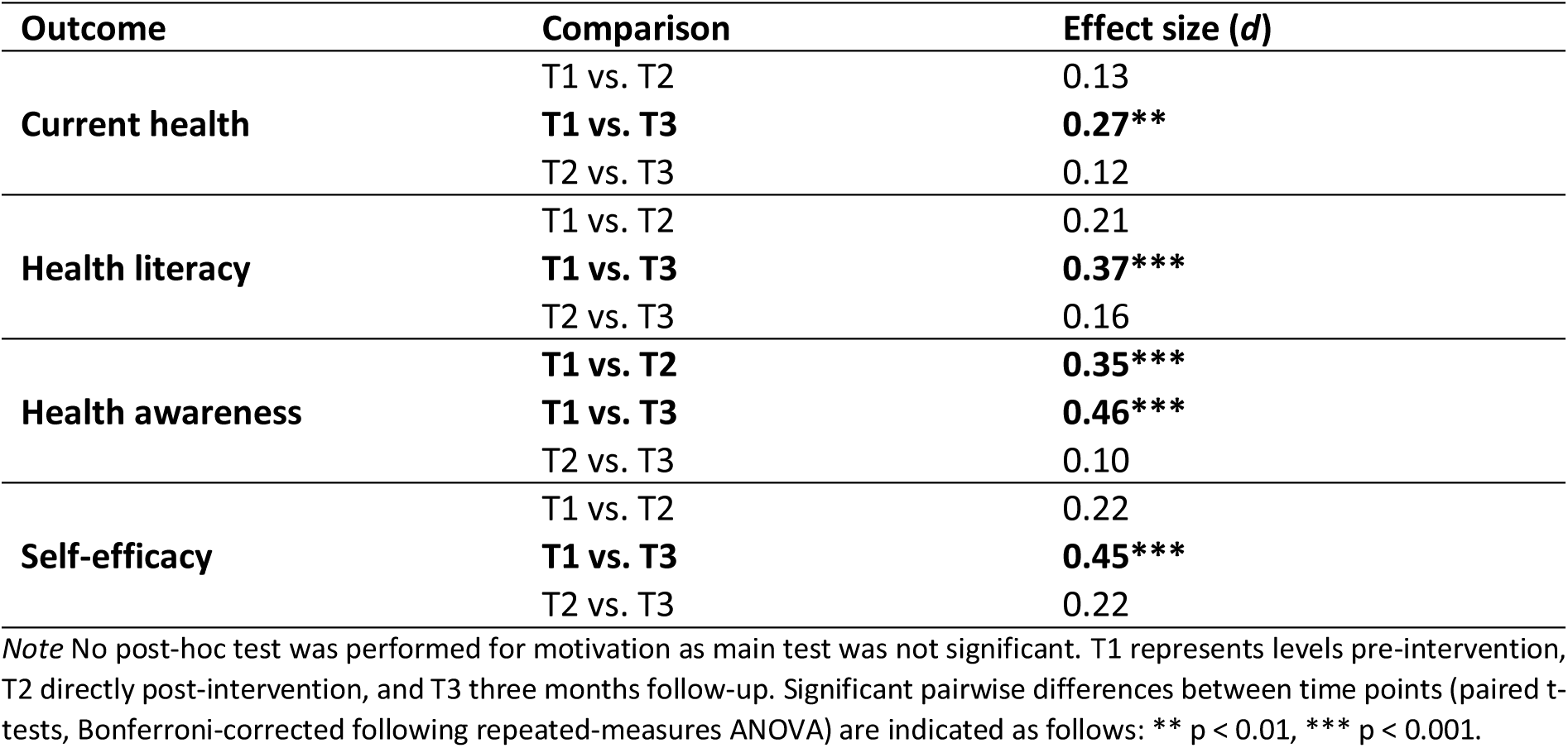
Effect sizes for the outcomes current health, health literacy, health awareness, and self-efficacy.

For all five outcomes, Mauchly’s test indicated a violation of the sphericity assumptions, which is why Greenhouse-Geisser-corrected degrees of freedom are reported. The repeated-measures ANOVA revealed a significant main effect of time on current health (*F*(1.81, 202.86) = 5.05, *p* = .009, η² = .011). Bonferroni-adjusted post hoc comparisons showed a significant difference between T1 and T3 (*t*(112) = 3.47, *p*adj = .002, *d* = 0.27), whereas the differences between T1 and T2 (*t*(112) = 1.41, *p*adj = .483, *d* = 0.13) and between T2 and T3 (*t*(112) = 1.70, *p*adj = .278, *d* = 0.12) were not significant. For health literacy, a significant main effect of time was detected (*F*(1.85, 206.88) = 8.865, *p* < .001, η² = .023). Post hoc comparisons showed a significant difference between T1 and T3, *t*(112) = 4.15, *p*adj < .001, *d* = 0.37. No significant difference was detected between T1 and T2 (*t*(112) = 2.23, *p*adj = .083, *d* = 0.21), as well as T2 and T3 (*t*(112) = 1.97, *p*adj = .153, *d* = 0.16). Health awareness showed a significant time effect as well (*F*(1.89, 212.1) = 17.25, *p* < .001, η² = .038). From T1 to T2 (*t*(112) = 4.05, *p*adj < .001, *d* = 0.35) as well as from T1 to T3 (*t*(112) = 5.44, *p*adj < .001, *d* = 0.46) showed significant differences in the post hoc analysis, while no significant difference was found from T2 to T3 (*t*(112) = 1.35, *p*adj = .537, *d* = 0.10). A significant main effect for time was found in self-efficacy as well (*F*(1.79, 200.99) = 11.639, *p* < .001, η² = .032). Post hoc test revealed significant differences from T1 to T3 (*t*(112) = 4.48, *p*adj < .001, *d* = 0.45) and from T2 to T3 (*t*(112) = 2.79, *p*adj = .019, *d* = 0.222), while no significant differences were found from T1 to T2 (*t*(112) = 2.35, *p*adj = .062, *d* = 0.22). The repeated-measures ANOVA revealed no significant main effect of time on motivation, *F*(1.75, 196.40) = 2.10, *p* = .131, η² = .006.

Friedman test was performed as a sensitivity analysis for the repeated measures ANOVA showing similar results in the main as well as post hoc tests. More detailed results on both analyses can be found in the Supplementary Material (Table S3 and S4).

## 4 Discussion

The primary aim of this prospective before-and-after observational study was to investigate whether the use of the PREVERY application for six weeks improved psychological capital and stress experience. In addition, questions were asked to assess participants’ motivation regarding their own health, their health literacy, and their health awareness, with the goal of identifying improvements in all of these areas. A significant time effect was observed for both the CPC and the PSS. The CPC score increased significantly from T1 to T2 and T3 and remained stable between T2 and T3, while the PSS score decreased markedly from T1 to T2, rose again by T3, and remained significantly below the baseline value; all pairwise comparisons were significant. The repeated-measures ANOVA also revealed significant time effects for self-reported current health, health literacy, health awareness, and self-efficacy (η² = .011–.038), with higher values at T3 than at T1 (for health awareness, this was already evident at T2; for self-efficacy, it was additionally evident between T2 and T3), but no time effect for motivation.

### 4.1 AI-based health coaching vs. human coaching

The results found in this study align with already existing literature. Several studies show the positive effects of digital health coaching on stress reduction (L’Engle et al., 2025; Lim et al., 2023; Wiegand et al., 2010) and increase in psychological well-being and capital (Peiper et al., 2025; Sacher et al., 2024). However, not all of them are AI-based interventions, but digital health coaching with therapists or hybrid formats using both AI and humans. A systematic review conducted by Loughnane et al. (2025) investigated different forms of digital health coaching in terms of trends, engagement, and lifestyle outcomes. Interestingly, they found that AI-based health coaching showed more consistent positive effects on physical activity levels, while human-based coaching led to stronger increases in psychological well-being and stress reduction (Loughnane et al., 2025). However, both AI and human digital coaching are acceptable and practical measures for supporting lifestyle changes, as the differences in outcomes between human and AI-based counseling systems were minimal (Loughnane et al., 2025).

AI health coaching has benefits as well as disadvantages over digital or analogous personal health coaching. The biggest advantage of AI coaches is their availability, as they can be consulted at any time and no foreplaning is necessary, whereas in-person coaching needs to be planned in advance and cannot be conducted spontaneously. However, there is a missing personal relationship between the AI coach and the patient. This personal relationship in in-person coaching is based on trust and empathy which might lead to long-term motivation (Loughnane et al., 2025). Nevertheless, the scalability of AI coaching might be better as more data can be processed objectively without personal interference.

### 4.2 Engagement

An important aspect in digital, but also analogous health coaching, seems to be engagement. Generally, higher and longer engagement leads to better health outcomes, goal attainment, increased retention, and adherence (Cueto et al., 2019; Loughnane et al., 2025). Loughnane et al. (2025) showed that digital but human coaching interventions caused higher retention and completion rates compared to AI and hybrid interventions. An analysis of the present data shows that most study dropouts were due to participants’ inability to meet the required weekly commitment. However, the guidelines for the present study were quite strict and failing to achieve the minimum score in even one of the six weeks resulted in exclusion from the study. A systematic review and meta-analysis investigated the dropout rates in app-based interventions for chronic disease (Meyerowitz-Katz et al., 2020). They found that the dropout rate for mHealth interventions is generally high. In observational studies in particular, the rate is even higher (49% dropout rate) than in randomized controlled trials (40% dropout rate) (Meyerowitz-Katz et al., 2020). These figures are in the same range as the current dropout rate of 55%, which was further driven by additional requirements.

It seems that the engagement is also driven by the length of intervention. Cueto et al. (2019) found that participants getting a 4- or 12-week health coaching intervention were less engaged than those participating in a 24-week trial. Within this context, it must be pointed out that the current study only consisted of six intervention weeks, further complicating the engagement of the participants.

Intriguingly, there have been significant changes in the follow-up period after the intervention finished. While the psychological capital and health awareness stayed on the same level compared to T2, showing the sustained effects of the intervention even after the end of the active phase, health literacy, current health, and self-efficacy became only significant in the follow-up period. However, although being decreased from T1 to T2, the perceived stress increased again from T2 to T3. Nevertheless, it seems that there are sustained effects of the intervention on some of the outcomes, especially self-efficacy due to potential consolidating the habits and goals established over the six weeks. One explanation could also be that around 59% of the participants used the app regularly (7%) or occasionally (52%) between T2 and T3 (Table S5). It must be noted that there might be expectation and confirmation biases, as well as response bias resulting from repeated completion of the survey driving this positive response in the follow-up period.

Within the present study, women were overrepresented compared to men, as they accounted for two-thirds of the study population. One reason for this disbalance could be the higher engagement rate of female participants in digital health coaching in general (Zainal et al., 2026). Therefore, male participants may not be able to reach the necessary growth points per week as easily as the female participants. However, this study found no difference in the sex distribution among those who dropped out and those who successfully completed the six weeks. Interestingly, Morgan et al. (2025) found that the usage of a mHealth app leads to reduction in blood pressure with higher reductions in the female compared to male participants. It, therefore, seems that stronger engagement rate in women and consequently higher usage time results in better health beneficial outcomes. This should be taken into account when interpreting the present findings as they might overestimate the actual effect due to the disbalance in sex distribution.

Existing research has shown that not only sex, but also age, is a main moderating factor in explaining the use of information and communication technologies like mHealth apps (Nunes et al., 2019). Nunes et al. (2019) found that performance expectancy, regardless of age, contributed significantly to the behavioral intention to use mHealth apps. However, effort expectancy was significantly moderated by sex and age. While older men use the app more often when they find it easy to use, younger men tend to use it less often under these circumstances. Furthermore, older women use mHealth offers more often when the usage has a positive influence on their social influence (Nunes et al., 2019). These results seem to partly explain the intentions behind the use of mHealth offers. However, it is difficult to draw conclusions on age-dependent usage intentions in the present analysis as the mean age is 47±13 years showing a relatively narrow distribution.

Although it is reported in the literature, that satisfaction is higher and more constant in studies incorporating human-facilitated coaching compared to AI-based health coaching interventions (Loughnane et al., 2025), the results of the current study show an increase in self-assessed self-efficacy, health awareness, and health behavior. One reason for the good results achieved with PREVERY could be its holistic approach, which combines AI-based health coaching with additional content and activities that promote personal health awareness and reinforce individual health behaviors. For example, some studies reported the limitations that the participants perceived some sort of repetitiveness, disconnection, and lack of warmth toward the AI chatbots (Aymerich-Franch & Ferrer, 2022; Ly et al., 2017; R. Williams et al., 2021).

It must be noted as well that participants with psychological conditions were excluded from the present study. They might represent the population with the highest need of stress reduction and self-efficacy increasing interventions. Their engagement rate may be much lower due to motivational issues and listlessness in for example depressed individuals (Smith, 2013). Therefore, the question remains if people with higher baseline stress benefit more or less from a health coaching program like PREVERY.

### 4.3 Practical relevance and biological mechanisms

One important aspect is the fact that statistical significance cannot be directly translated into practical relevance. Especially when investigating stress scales, an improvement on these scales must not lead to a noticeable reduction in stress experience (Reis et al., 2019). Therefore, the present study not only assessed stress via the PSS and CPC, but also asked for the participant’s subjective perception of their self-efficacy. Interestingly, this perception of self-efficacy as well as their subjective health awareness and competency was significantly improved from start of the intervention to the end (six weeks) and also three months later. Even though the practical relevance of the reduction in PSS seems questionable at first, it is indeed supported by the significant improvements in health awareness, self-efficacy, and health literacy.

Although there is a psychological measure of the stress reduction and perception in the present study, an additional perspective could be given by related biomarkers such as interleukin (IL)-6, cortisol, or C-reactive protein (CRP) assessing the actual biological stress and inflammatory response. This has been poorly studied so far and gains increasing attention only now (Wang & Zhong, 2026). A systematic review and meta-analysis conducted by O’Toole et al. (2018) examining 19 studies found a significant reduction in CRP following psychological interventions. However, none of them investigated digital health coaching. Interestingly, Lozano-Lozano et al. (2019) investigated the effects of mHealth app usage on inflammation-related biomarkers in breast cancer survivors. They showed, that their intervention reduced CRP and IL-6 significantly. It should be noted, however, that these authors, just like the studies included in the meta-analysis mentioned above, did not conduct any interventions comparable to the PREVERY health coaching app in this study. Therefore, future studies should investigate, which underlying biological pathways lead to the reduction in stress perception and if a biological mechanism can be found.

Terblanche (2020) developed the Designing AI Coach (DAIC) framework which allows a structured assessment of AI standards in health coaching. There are four principals that should be incorporated in AI-based health coaching. The first is building a relationship that is based on empathy, trust, and transparency. Second, the concept on which the AI health coaching is build must be based on an existing theoretical framework and, third, ethical standards protecting data security, data transparency, clear responsibilities, preserving the user’s autonomy, and avoiding algorithmic bias. Lastly, the AI coach must have a clearly defined focus, concentrating on, for example, goal setting, well-being, self-efficacy, or behavioral change (Terblanche, 2020).

PREVERY meets criteria one, two, and four. The first criterion is met through prompts and the app’s design, which are managed by health experts, as well as through the labeling of AI-generated and other evidence-based content. The second criterion is met through strict adherence to the HAPA and Tiny Habits models, while the fourth is met by the app’s architecture itself, since no standalone chatbot is used. Instead, a consistently defined prevention process was built, and only at specific points the pre-sorted user data was sent to the LLM along with the corresponding prompts. The third criterion was only met after the study concluded through further development of the app. The app now uses more EU-based services instead of U.S.-based ones. Interestingly, the studies identified by Loughnane et al. (2025) adapted all the first two domains, but only a few studies addressed the ethical component and all but two studies defined a clear focus of their AI coach.

### 4.4 Limitations and perspectives

The present study results must be interpreted within their respective limitations. First, the single-group before-and-after design without a control group does not allow to draw causal conclusions. Changes in perceived stress, psychological capital, and self-efficacy could, therefore, be partly explained by factors unrelated to the intervention. These include regression to the mean, particularly if individuals with higher stress levels were more likely to enroll in the study, as well as natural fluctuations in stress levels over time. Furthermore, all outcomes were self-reported, so expectation and social desirability effects cannot be ruled out completely. Finally, participants that voluntarily enroll in a digital prevention program are likely to have higher motivation than the general population, which limits the generalizability of the findings. A randomized controlled trial with a waiting list or an active control group is needed to support the observed effects.

Second, for the analysis, participants had to meet a defined minimum usage requirement: at least 120 growth points within six weeks and at least 20 points per week. This requirement was based, among other things, on criteria related to a potential future ZPP certification. At the same time, this may have limited the generalizability of the results, as individuals with lower usage intensity were not included in the analysis. It is not possible to clearly assess the extent to which these factors contributed to the decline from 253 registrations to 113 participants in the active study phase. For future studies, it would, therefore, be interesting to systematically document the reasons for non-participation or dropout, as well as differences in usage intensity.

Third, the per-protocol analysis provides insights into outcomes among participants meeting the predefined intervention exposure criteria. However, these findings may not be generalizable to users with lower engagement levels, and selection bias may have influenced the observed effect sizes. The effects sizes should, therefore, be interpreted in the light of this limitation.

The fourth point concerns the technical design of the study version of PREVERY. Although the web app was optimized more for desktop use, many of the respondents used PREVERY primarily on their smartphones. This may have affected ease of use, particularly navigation and the completion of tasks and goals. In addition, there were no integrated push notifications, so reminders were sent via email, among other methods. The qualitative feedback, as well as requests for more concrete weekly plans and mini-exercises relevant to daily life, also suggest that, in addition to the personalization of health information, there is room to provide stronger support for the concrete implementation of personal goals in everyday life. For future versions, it would, therefore, be interesting to investigate to what extent a stronger focus on mobile use, as well as low-threshold exercises, planning, and reminder functions, can help improve the connection between individual health goals and their implementation in daily life.

Given the above-mentioned limitations, a randomized controlled trial should be performed in the future, investigating the effects of the app usage to a control group, assessing not only psychological variables, but actual stress- and inflammation-related biomarkers. A longer follow-up period is needed to verify the long-term effects of the intervention. In the best case, the app usage only would be compared to hybrid or in-person interventions to enable a direct comparison between different health coaching methods.

The study results and qualitative feedback should be interpreted in light of the fact that the version of the program examined in the study does not reflect PREVERY’s current state of development. Since the study was completed, the application has been further developed, particularly with regard to mobile usability and providing practical, everyday support for achieving goals.

## 5 Conclusion

In this prospective before-and-after observational single-arm study, the use of PREVERY over a six-week period was associated with a reduction in perceived stress as well as an increase in psychological capital, self-efficacy, health awareness, and health literacy, which persisted even at follow-up. Therefore, PREVERY seems to be a prevention program that initiates stress-reducing processes and contributes to an improved perception of one’s own mental health. Overall, PREVERY offers a cost-effective and highly scalable solution for health coaching, as it enables continuous support compared to expensive analog options. Given the absence of a control group and relevant stress-related biomarkers, future research should validate these positive and health beneficial findings within a randomized controlled trial.

## Data Availability

All data produced in the present study are available upon reasonable request to the authors.

## 6 Acknowledgments

We thank the BERGISCHE KRANKENKASSE for their support in participant recruitment.

## 7 Statements and declarations

### 7.1 Informed Consent

Informed consent was obtained for any experimentation with human subjects.

### 7.4 Data availability statement

The data will be provided upon justified request.

### 7.5 Author contributions

**MR**: Formal Analysis, Visualization, Writing – original draft. **TT**: Conceptualization, Methodology, Writing – review & editing. **TT**: Conceptualization, Methodology, Writing – review & editing. **WB**: Conceptualization, Supervision, Methodology, Writing – review & editing.

